# Obesity and accelerated epigenetic aging in a high-risk cohort of children

**DOI:** 10.1101/2021.11.03.21265865

**Authors:** Laura Etzel, Waylon J. Hastings, Molly A. Hall, Christine Heim, Michael J. Meaney, Jennie G. Noll, Kieran J. O’Donnell, Irina Pokhvisneva, Emma Jane Rose, Hannah M. C. Schreier, Chad Shenk, Idan Shalev

**Affiliations:** Department of Biobehavioral Health, The Pennsylvania State University, University Park, PA, USA; Department of Veterinary and Biomedical Sciences, The Pennsylvania State University, University Park, PA, USA; Charité – Universitätsmedizin Berlin, corporate member of Freie Universität Berlin, and Humboldt-Universität zu Berlin, Berlin Institute of Health (BIH), Institute of Medical Psychology, Berlin, Germany; Departments of Psychiatry and Neurology and Neurosurgery, McGill University, Montreal, Québec, Canada; Department of Human Development and Family Studies, The Pennsylvania State University, University Park, PA, USA; Yale Child Study Center; Department of Obstetrics Gynecology and Reproductive Sciences, Yale School of Medicine, Yale University, New Haven, CT, USA; Ludmer Centre for Neuroinformatics and Mental Health, Douglas Mental Health University Institute, McGill University, Montreal, QC, Canada; Edna Bennett Pierce Prevention Research Center, The Pennsylvania State University, University Park, PA, USA; Department of Pediatrics, The Pennsylvania State University College of Medicine, Hershey, PA, USA

**Keywords:** Obesity, Accelerated Aging, Epigenetic Age, Pace of Aging, Childhood

## Abstract

**Background:** New insights into mechanisms linking obesity to poor health outcomes suggest a role for cellular aging pathways, casting obesity as a disease of accelerated biological aging. Although obesity has been linked to accelerated epigenetic aging in middle-aged adults, the impact during childhood remains unclear. We tested the association between body mass index (BMI) and accelerated epigenetic aging in a cohort of high-risk children. Participants were children (N=273, aged 8 to 14 years, 82% investigated for maltreatment) recruited to the Child Health Study, an ongoing prospective study of youth investigated for maltreatment and a comparison youth. BMI was measured as a continuous variable. Accelerated epigenetic aging of blood leukocytes was defined as the age-adjusted residuals of several established epigenetic aging clocks (Horvath, Hannum, GrimAge, PhenoAge) along with a newer algorithm, the DunedinPoAm, developed to quantify the pace-of-aging. Hypotheses were tested with generalized linear models.

**Results:** Higher BMI was significantly correlated with older chronological age, maltreatment status, household income, blood cell counts, and three of the accelerated epigenetic aging measures: GrimAge (r=0.29, *P*<.*0001*), PhenoAge (r=0.25, *P*<.*0001*), and DunedinPoAm (r=0.37, *P*<.*0001*). In fully adjusted models, GrimAge (b=.06; *P=*.*007*) and DunedinPoAm (b=.0017; *P*<.*0001*) remained significantly associated with higher BMI. Maltreatment-status was not independently associated with accelerated epigenetic aging after accounting for other factors.

**Conclusion:** In a high-risk cohort of children, higher BMI predicted epigenetic aging as assessed by two epigenetic aging clocks. These results suggest the association between obesity and accelerated epigenetic aging begins in early life, with implications for future morbidity and mortality risk.

## 1.1 Background

Obesity remains a pressing global health issue, despite decades of attention and research. Comorbidities associated with obesity overlap with aging and age-related phenotypes, and it is hypothesized that obesity may accelerate aging across a range of cellular and bodily systems, leading to increased risk of age-related diseases.^1–3^ The health impacts of obesity may also begin in childhood, as obese children face increased risk of childhood onset psychological and physical morbidities (e.g., depression, systemic inflammation, type II diabetes, cardiovascular abnormalities).^4^ Given the profound public health burden of obesity, understanding the early-life etiology of the association between obesity and accelerated biological aging is critical for the mitigation of future disease risk.

Several approaches for quantifying biological aging have been used to assess links with obesity, including telomere length and epigenetic aging clocks.^5–8^ Accelerated epigenetic aging, or the difference between an individual’s epigenetic age and their chronological age, has been independently associated with both obesity and early morbidity and may be an indicator linking obesity with early onset of diseases of aging.^9^ Regulation of methylation at many cytosine-phosphate-guanine dinucleotides (CpGs) is positively correlated with chronological age, a finding that led to the creation of first-generation epigenetic algorithms (i.e., the Horvath and Hannum clocks), which use methylation levels to predict chronological age.^10,11^ A second generation of epigenetic clocks, built on physiological measures of health and disease risk, aimed to predict lifespan and healthspan (i.e., GrimAge and PhenoAge clocks).^12,13^ Most recently, a new generation epigenetic clock predicting the rate of biological aging, the Dunedin Pace of Aging methylation clock (DunedinPoAm) was introduced using longitudinal data on organ-system integrity across 12 years.^14^ The DunedinPoAm aims to quantify the pace of an individual’s biological aging through a single-time-point measure of DNA methylation.

Prior work has linked accelerated epigenetic aging with obesity. Horvath et al.^7^ demonstrated obesity accelerates epigenetic aging in the liver, which resulted in approximately 2.7 years age acceleration for every 10-point increase in body mass index (BMI). Using the same Horvath clock, another study linked accelerated epigenetic aging in adipose tissue to obesity status.^15^ In a longitudinal study, an increase in BMI was significantly related to accelerated epigenetic aging in white blood cells of middle-aged adults.^8^ Similarly, in a large sample of adult women, epigenetic age acceleration in blood was associated with higher BMI, larger waist circumference, and greater waist-to-hip ratio using first- and second-generation clocks (i.e., Horvath, Hannum, PhenoAge, and GrimAge).^16^ Although work in relation to epigenetic aging and obesity in children is scarce, a recent study looking at socioeconomic disadvantage in children reported a correlation between higher BMI and faster salivary DunedinPoAm-measured pace of biological aging.^17^

Early-life adversity is a potent risk factor for childhood obesity. Longitudinal and cross-sectional studies demonstrate exposure to childhood maltreatment (CM; e.g., physical/sexual abuse and neglect) is associated with accelerated increases in BMI through adolescence and early adulthood, thereby elevating risk of obesity in adulthood.^18–20^ Differences in epigenetic programming offer an intriguing possibility to further account for this association. Developing an understanding of the association between obesity and accelerated aging in early life, both in the moderating context of CM and more broadly in all children, is critical to prevention efforts of many obesity-related negative health outcomes. Our study is the first to comprehensively examine the association between accelerated epigenetic aging and BMI in children using multiple epigenetic clocks. Using data from the Child Health Study (CHS), a cohort study of children with a high prevalence of Child Protective Services investigations due to suspected CM exposure, we tested the association between BMI and accelerated aging in childhood using a diverse panel of first-, second-, and new-generation epigenetic clocks. We hypothesized that BMI would be associated with accelerated epigenetic aging. Given the high prevalence of reported maltreatment in our cohort, we additionally explored the role of CM in this association.

## 1.2 Results

### 1.2.1 Sample Descriptives

No significant differences were detected in mean age, distribution of biological sex, or ethnicity between the maltreatment and comparison groups. For both the full cohort (N=439) and our analytic sample (N=273), the maltreatment group was lower income (maltreatment mean=$33,000 [SD $32,000], comparison mean=$54,000 [SD $39,000]; *P=*.*009*) and had a higher BMI (maltreatment mean=22.1 [SD 6.1], comparison mean=20.3 [SD 5.3]; *P=*.*04*) (see Additional File Table S1 for full cohort sample demographics). There were additional group differences in ‘Other’ racial identification (Table 1). Family income was the only demographic factor to remain significant after correction for multiple comparisons at 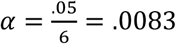. Correlations among the outcomes and covariates are depicted in Table 2.

**Table 1.** Summary Demographics for Analytic Sample

| Variable, mean (SD) | Total (N=273) | Maltreatment (N=225) | Comparison (N=48) | P-value |
| --- | --- | --- | --- | --- |
| Body Mass Index (BMI) | 21.8(6.0) | 22.1(6.1) | 20.3(5.3) | .04* |
| Age (at collection) | 11.4(1.5) | 11.4(1.5) | 11.1(1.5) | .15 |
| Biological Sex M(F)% | 49.5%(50.5) | 49.3%(50.7) | 50.0%(50.0) | .93 |
| Income \$10,000/year | 3.7(3.5) | 3.3(3.2) | 5.7(4.0) | .0003** |
| Race |  |  |  |  |
| Black | 16.5% | 16.9% | 14.6% | .70 |
| White | 67.8% | 65.3% | 79.2% | .06 |
| Other | 15.8% | 17.8% | 6.3% | .04* |
| Ethnicity |  |  |  |  |
| Hispanic | 11.4% | 12.9% | 4.2% | .08 |

†P<.10, \*P<.05, \*\*P<.01, \*\*\*P<.0001

**Table 2.** Correlations among Outcomes and Covariates

|  | BMI | Age | Sex (Male) | Maltreatment | Household Income | Proportion Lymphocytes | Proportion Monocytes | Proportion Granulocytes | HorvathAA | HannumAA | GrimAgeAA | PhenoAgeAA |
| --- | --- | --- | --- | --- | --- | --- | --- | --- | --- | --- | --- | --- |
| Age | 0.29*** | -- |  | -- | -- | -- | -- | -- | -- | -- | -- | -- |
| Sex (Male) | -0.12 | -0.07 | -- | -- | -- | -- | -- | -- | -- | -- | -- | -- |
| Maltreatment | 0.11† | 0.09 | -0.010 | -- | -- | -- | -- | -- | -- | -- | -- | -- |
| Household Income | -0.18** | -0.005 | 0.04 | -0.26*** | -- | -- | -- | -- | -- | -- | -- | -- |
| Proportion Lymphocytes | -0.28*** | -0.06 | 0.14* | -0.01 | 0.10 | -- | -- | -- | -- | -- | -- | -- |
| Proportion Monocytes | 0.13* | 0.07 | 0.04 | -0.06 | -0.03 | -0.48*** | -- | -- | -- | -- | -- | -- |
| Proportion Granulocytes | 0.27*** | 0.05 | -0.17** | -0.03 | -0.11† | -0.97*** | 0.27*** | -- | -- | -- | -- | -- |
| HorvathAA | 0.04 | 0.01 | 0.06 | 0.02 | -0.04 | -0.01 | -0.07 | 0.02 | -- | -- | -- | -- |
| HannumAA | 0.12† | 0.01 | -0.08 | -0.03 | -0.05 | -0.58*** | 0.35*** | 0.53*** | 0.30*** | -- | -- | -- |
| GrimAgeAA | 0.29*** | 0.002 | 0.12† | 0.14* | -0.32*** | -0.48*** | 0.28*** | 0.45*** | 0.08 | 0.40*** | -- | -- |
| PhenoAgeAA | 0.25*** | 0.01 | -0.22** | 0.03 | -0.09 | -0.71*** | 0.33*** | 0.69*** | 0.27*** | 0.71*** | 0.48*** | -- |
| Dunedin PoAm | 0.37*** | -0.02 | -0.15* | 0.08 | -0.13* | -0.68*** | 0.31*** | 0.67*** | 0.09 | 0.54*** | 0.54*** | 0.67*** |

† $P < .10$ , \* $P < .05$ , \*\* $P < .01$ , \*\*\* $P < .0001$

### 1.2.2 BMI and epigenetic aging clocks

BMI was moderately correlated with age (r=0.29, *P*<.*0001*), household income (r=-0.18, *P=*.*004*), proportion lymphocytes (r=-0.28, *P*<.*0001*), proportion granulocytes (r=0.27, *P*<.*0001*), and three epigenetic age acceleration clocks: GrimAge (r=0.29, *P*<.*0001*), PhenoAge (r=0.25, *P*<.*0001*), and DunedinPoAm (r=0.37, *P*<.*0001*). Excepting the correlation between BMI and income, these remained significant after correction for multiple comparisons at 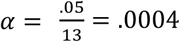 The correlation between BMI and the HannumAA did not reach significance (r=0.12, *P=*.*055*) and BMI was not correlated with the HorvathAA (r=0.04, *P=*.*46*).

In models adjusting for sex, race, ethnicity, and family income, higher BMI was consistently associated with GrimAge acceleration and PhenoAge acceleration (see Additional File Tables S2-S6 for full model specifications). After accounting for blood cell proportions, BMI remained significantly associated with accelerated aging measured by GrimAge (b=.06; CI: 0.02, 0.10; *P=*.*007*) but was attenuated below α=.05 for PhenoAge acceleration (b=.05; CI: − 0.03, 0.13; *P=*.*25*). A BMI increase of one standard deviation above the cohort mean was associated with an additional 0.42 years of accelerated aging as measured by GrimAge (Figure 1). BMI was not associated epigenetic age acceleration derived from the Horvath or Hannum clocks in any model tested (all *P*>.*05*).

**Figure 1.**
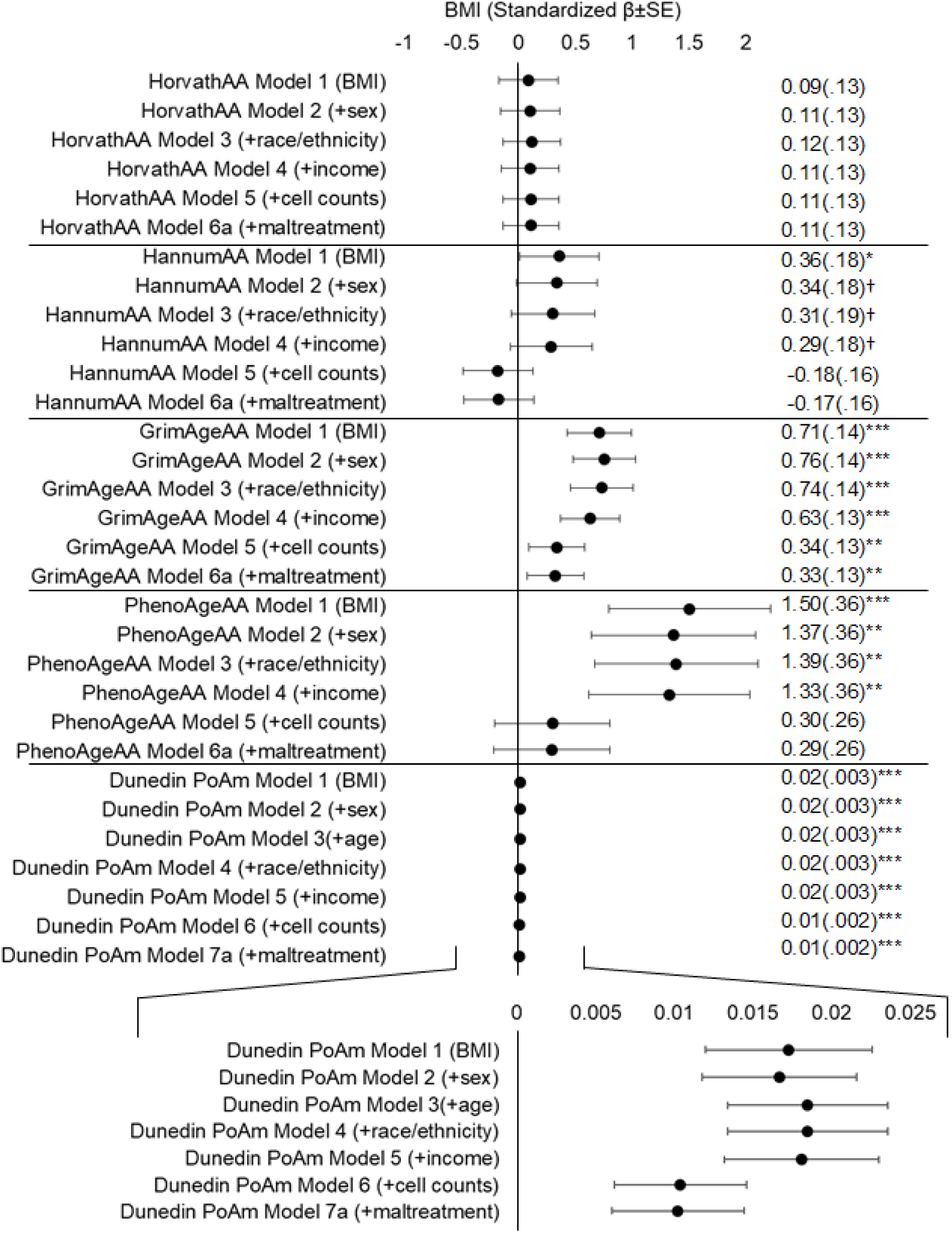
Standardized β+SE estimates for BMI across all accelerated epigenetic aging and DunedinPoAm outcomes. Each row specifies the stepwise addition of a covariate to the base model of accelerated epigenetic aging outcome regressed onto BMI. Zoomed-in view of estimates for models for DunedinPoAm. ^*†*^*P*<.*10, *P*<.*05, **P*<.*01, ***P*<.*0001*

Higher BMI was significantly associated with a faster pace-of-aging as measured by the DunedinPoAm (b=.0017; CI: 0.001, 0.002; *P*<.*0001*) adjusting for all covariates. A BMI increase of one standard deviation above the cohort mean was associated with an increase of 0.012 years in the pace of epigenetic aging. Associations between BMI and accelerated aging measured by GrimAge and DunedinPoAm remained significant after correcting for multiple comparisons at 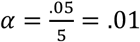.

### 1.2.3 Maltreatment exposure, BMI, and epigenetic aging clocks

Given the opportunity to explore accelerated epigenetic aging in this cohort of children with a high prevalence of maltreatment exposure, we tested whether maltreatment was associated with accelerated epigenetic aging or moderated the association between BMI and accelerated epigenetic aging. Though BMI was significantly different between the maltreatment and comparison groups in initial demographic comparisons (maltreatment BMI mean=22.1 (SD 6.1), comparison BMI mean= 20.3 (SD 5.3); *P=*.*04*), this difference was attenuated to non-significance with the inclusion of covariates (see Additional File Table S7 for full model specifications). Maltreatment was not independently associated with epigenetic age acceleration in our analytic sample and did not moderate the association between BMI and accelerated epigenetic aging measures.

## 1.3 Discussion

In a cohort of high-risk children, we tested the association between BMI and accelerated aging using three generations of epigenetic aging measures. We further explored whether maltreatment status moderated this association. Higher BMI in childhood was associated with accelerated epigenetic aging and accelerated pace of epigenetic aging after accounting for covariates. Specifically, children with a higher BMI had an accelerated GrimAge, an epigenetic aging measure designed to predict time to death that also associates with cardiovascular disease risk.^13^ Children with higher BMI also demonstrated an accelerated pace of epigenetic aging using the DunedinPoAm, a DNA methylation clock built by measuring organ-system decline from young adulthood to midlife.^14^ Recent work in a cohort of socioeconomically disadvantaged children observed a correlation between BMI and DunedinPoAm-measured pace of aging in saliva DNA,^17^ a finding that is supported and extended here in leukocyte DNA through robustly controlling for additional covariates known to influence both BMI and measures of epigenetic aging. Previous work has found an association between obesity and accelerated epigenetic aging using the Horvath clock in adult liver tissue^7^ and in whole blood of middle-aged adults.^8^ Our results lend support for the development of this association in childhood.

Exploration of the relation between maltreatment and accelerated epigenetic aging failed to replicate previous work in this domain, though important differences exist between study designs. For example, recent studies in children using salivary DNA to calculate HorvathAA found that threat-based, but not deprivation-based, maltreatment was related to accelerated epigenetic aging,^21^ and that direct exposure to violence accelerated epigenetic aging.^22^ Data collection for the CHS is ongoing, thus we are exploring maltreatment association in a subset of the final cohort using a dichotomous maltreatment status variable (‘yes’, ‘no’). The currently restricted sample and binary categorization of maltreatment limits our ability to dissect associations between specific types of maltreatment and accelerated epigenetic aging that may exist in this high-risk sample. It is plausible that experiences of violence and threat within the CHS cohort may have unique effects on epigenetic aging that may be revealed with future collection of detailed knowledge around the types of maltreatment exposure for participants.

Higher BMI across the lifespan has been associated with early onset of age-related diseases and mortality, generating interest in the link between obesity and aging.^23^ Although we observed an association between obesity and accelerated epigenetic aging that is already present in childhood, the cross-sectional nature of our data prevents us from addressing mechanism and causality. Longitudinal work in the Avon Longitudinal Study of Parents and Children provides early support for obesity as a driver of variation in DNA methylation.^24^ Mechanistic links connecting obesity and accelerated aging have yet to be fully elucidated. Research suggests, however, that cellular processes of fat storage, inflammation, oxidative stress, and energy homeostasis may be involved.^2,25,26^ Dysregulation in these processes may drive epigenetic alterations resulting in an accelerated epigenetic aging profile. Methylation of CpG sites is a plastic process moderated by multifactorial intrinsic and extrinsic processes. Obesity has been linked to both hyper- and hypo-methylation of CpG sites across the genome, which in turn associate with the regulation of leptin, adiponectin, and other features of cellular energy balance.^24,27,28^ Direct evidence of obesogenic factors altering methylation patterns related to aging is lacking and support for this framework derives mainly from evidence of the impact nutrition and caloric restriction may have in altering epigenetic aging.^9,29,30^

The prevalence of childhood obesity has increased dramatically over the past four decades.^31^ Our findings of accelerated epigenetic aging and an accelerated pace of epigenetic aging in children with higher BMI have important implications for health and the mitigation of future disease risk. Children with obesity are at increased risk for childhood and early-onset adult diseases. Chronic conditions, such as type 2 diabetes, hypertension, atherosclerosis, and a range of other negative mental and physical health outcomes are more prevalent in individuals with childhood obesity.^32–35^ These diseases occur often, raising the cost of medical care and further increasing the likelihood of early-onset disability and a reduced lifespan. Many of the diseases associated with childhood obesity are also diseases of aging. Our findings point to the possible role of accelerated aging at the cellular level in driving the early-onset disease phenotypes seen in children with obesity as they become adolescents and adults.

We acknowledge several limitations. Recruitment of participants, in particular demographically matched comparison children, is ongoing. Although the findings of accelerated epigenetic aging and accelerated pace of epigenetic aging do not hinge on additional recruitment within the CHS cohort, the exploratory analysis of maltreatment as a modifier of the obesity-epigenetic aging association should be understood as preliminary and used as a direction for future cross-sectional and longitudinal research with the final assembled cohort. Along these lines, future analyses of data from this cohort will be able to include more nuanced maltreatment variables including duration of abuse, age of abuse onset, type of abuse, and polyvictimization status. The addition of these variables will be critical to understanding the associations between both maltreatment and obesity, as well as maltreatment and accelerated aging within this cohort. For example, timing of abuse has been shown to be a critical factor in the association between CM and epigenetic aging.^36^ Further, additional consideration should be given to the generalizability of these findings within low-risk cohorts of children.

The use of BMI as a variable to conceptualize body fat mass, although widely used, is also contended. Evidence exists that in both childhood and adulthood, the specific areas of fat mass accumulation (e.g., upper body, lower body, subcutaneous, visceral) may be critical to determining future disease risk,^37,38^ an issue that may be particularly relevant for the deposition of fat following maltreatment.^39^ Using BMI instead of more nuanced measures of adiposity distribution may obscure associations among accelerated epigenetic aging, obesity, and maltreatment within our current sample of children. Finally, mechanistic understanding of the association between biological processes of aging and the three generations of epigenetic clocks examined here is lacking. These clocks are derived from regressing observed features of aging, morbidity, and mortality onto measured epigenetic changes in adulthood. These correlations do not necessarily yield direct insights into the processes of biological aging. Investigations into CpGs of some epigenetic clocks demonstrate enrichment for genes involved in organismal development and cell survival,^11^ and a tendency to colocalize within glucocorticoid response elements.^40^ However, it remains uncertain whether this is the case for all epigenetic clocks, which are constructed using a variety of CpGs. The relative contribution of individual CpGs is also obscured when tens or hundreds are collapsed into a single composite measure. Despite this limitation, these epigenetic aging measures may have important clinical and research utility in obese pediatric populations.

## 1.4 Conclusions

Our findings point to the possible utility of epigenetic aging measures for future clinical and research work in children with obesity. Epigenetic aging and pace of epigenetic aging measures may serve as useful proxies when assessing the efficacy of obesity interventions for children.^41,42^ The true endpoints for obesity interventions, risk of obesity-related diseases and reduced lifespan, often take years to decades to manifest. These timescales often preclude longitudinal intervention studies from using disease risk and lifespan as outcome measures due to cost and complexity of study design. Epigenetic aging and pace of epigenetic aging measures may be appropriate outcomes by which interventions can be judged as efficacious in ameliorating childhood obesity-related disease risks.

## 1.5 Materials and Methods

### 1.5.1 Study Design and Sample Recruitment

Participants for this study were drawn from the ongoing CHS, a large multidisciplinary study designed to provide prospective, longitudinal data on the health and development of children with and without a history of maltreatment. The CHS is recruiting a large state-wide cohort of children recently investigated for CM and non-maltreated comparison children. The goals of the CHS are to elucidate the multiple etiological processes, as well as mediators and moderators, believed to play a role in the onset and maintenance of adverse health outcomes among victims, and to better inform intervention opportunities to reverse the negative consequences of maltreatment.

Children with a recent (<12 months) report of maltreatment exposure are identified in collaboration with Pennsylvania’s Statewide Child Welfare Information System (CWIS). Subjects with recent involvement in the CWIS are invited to participate in the study through home mailings and phone contact by study coordinators. Eligibility criteria includes: (1) aged 8 to 13 years, (2) subject of a CWIS maltreatment report (i.e., an allegation is made and investigated) and agreement for participation within 12 months of CWIS involvement, and (3) agreement of participation by a non-abusing caregiver. Non-maltreated comparison children are recruited via targeted advertisements from the same Pennsylvania counties as maltreated children with the goal of demographically matching at least one maltreated child based on age, race, ethnicity, sex, income level, and region within the State. Eligibility for participation includes: (1) no previous CWIS reports or contact, and (2) demographic match to a maltreatment participant. After recruitment, participating families are invited to visit the Center for Healthy Children at The Pennsylvania State University. The Center was established to serve as a dedicated on-campus facility for the CHS. The physical space houses a research lab with areas dedicated to biospecimen sample collection, measurement of physical health by trained nursing staff, participant and family interview rooms, and physical/hard-copy data storage.^43^ The Pennsylvania State University Institutional Review Board approved the study, and informed assent (child) and consent (caregiver) was obtained from all participants.

Cross-sectional data reported here were drawn from the baseline (i.e., Time 1) assessment of currently enrolled CHS participants. Although recruitment for this cohort study is ongoing with a target enrollment of 900 children, an initial subset of 439 was available for the purposes of these analyses. Of the 439 participants who have completed Time 1, 435 consented to anthropometric measurements and 401 consented to and successfully completed blood draws (1 caregiver refusal, 33 participant refusals, 4 attempted but incomplete blood draws). The first 300 participant identification numbers were sent as the first batch for methylation analysis (constituting 286 total samples sent) and 273 samples survived methylation QC measures. These 273 participants are our final analytic sample. Summary statistics for participants included in our analyses are provided in Table 1 (see Additional File for comparative summary statistics of the 439 participants available at Time 1).

### 1.5.2 Body Mass Index

Anthropometric surveys, including height and weight, were conducted by trained CHS staff during participants’ visit to the research lab. BMI was defined as total body mass in kilograms divided by the squared body height. As a sensitivity analysis, we tested a dichotomous obesity variable (obese vs. non-obese) as determined by age- and sex-specific BMI > 95^th^ percentile cutoffs using the Center for Disease Control’s Childhood BMI Calculator.^44^ Results were unchanged using a dichotomous obesity variable and we report results below using only the continuous BMI variable.

### 1.5.3 Assessment of DNA Methylation and Calculation of Accelerated Aging Variables

Genomic DNA was extracted from whole blood using a semi-automated approach (Qiasymphony, Qiagen). Genomic DNA purity and concentration was assessed using a nanophotometer (ImplenP300, Implen). Infinium methylation EPIC Beadchip (EPIC array, Illumina, San Diego CA, USA) was used to describe variation in DNA methylation across the genome. Genomic DNA (1ug) from whole blood was treated with sodium bisulfite using the Zymo EZ-96 DNA Methylation Kit™ (Zymo Research, Orange, CA, USA) with 200ng of bisulfite-treated DNA amplified, fragmented, and hybridized on the EPIC array. Samples were randomized across plates to avoid potential confounding between sources of technical variation and phenotypes of interest (e.g., maltreatment status). The resulting raw intensity values (idat files) are directly loaded into R for quality control and normalization using the Meffil package.^45^ We used normal-exponential out-of-band (noob) for background correction and dye-bias adjustment. Samples and probes with low signal intensity were removed. Concordance between predicted biological sex based on DNA methylation data and self-reported gender were verified for each sample with discordant samples removed. Finally, we used a Bayes method (ComBat) to correct for sources of technical variation (i.e., slide).^46^ Resulting methylation levels were used to calculate five separate measures of epigenetic age: the Horvath clock, Hannum clock, GrimAge, PhenoAge, and the DunedinPoAm. Measures of epigenetic age acceleration (HorvathAA, HannumAA, GrimAgeAA and PhenogeAA) were derived from beta values using a publicly available tool.^47^ Likewise, the Dunedin methylation pace of aging (DunedinPoA) score was derived from the DNA methylation of 46 CpGs as described previously.^14^

### 1.5.4 Other Measures

Income, biological sex, race, ethnicity, and predicted blood cell proportions (described using DNA methylation data and a reference-based approach)^48^ were included as covariates due to known associations with BMI and epigenetic aging measures. Income was assessed via caregiver self-report as current total household family income before taxes in increments of $10,000 (e.g., under $10,000 coded as ‘0’, $10,000-$19,999 coded as ‘1’, $20,000 - $29,999 coded as ‘2’ and an income over $120,000 coded as ‘11’). Biological sex was determined via self-report as either ‘male’ or ‘female’ and cross-validated using the DNA methylation predicted sex. Two participants self-identified as ‘other/transgender’ but had not undergone any gender-reassignment treatments and were therefore coded as their cross-validated biological sex. Race and ethnicity were reported by caregivers for participating youth. Race was coded as ‘White/Caucasian’, ‘Black/African American’, or ‘Other’ (American Indian, Alaskan Native, Asian, Pacific Islander, Multiracial, or Other). Ethnicity was reported as either ‘Hispanic’ or ‘Non-Hispanic’. Proportions of lymphocytes (summed estimates for CD8, CD4, natural killer, and B cells), monocytes, and granulocytes were extracted from epigenetic estimates of blood cell counts on the same blood used for methylation measures using an established reference-based approach, and included as covariates as needed for certain robustness checks.^48^

Given the association between BMI and pubertal stage,^49^ we additionally investigated pubertal stage as a covariate. Pubertal stage was assessed using Tanner staging, an index of physical ratings from 1 (prepubertal), 2 (pubertal onset, presence of breast buds and pubic hair) through 5 (fully mature) with each participant giving two separate ratings (breast/testis development and pubic hair development).^50,51^ A final pubertal stage was conceptualized as the average of these two measures of pubertal development. The correlation between BMI and pubertal stage (r=0.34, *P*<.*0001*) was similar in size, direction, and strength to that of BMI and age, and the correlation between pubertal stage and age was moderate and positive (r=0.65, *P*<.*0001*). Correlations between pubertal stage and accelerated epigenetic aging variables were non-significant with the exception of GrimAgeAA (r=0.18, *P=*.*002*). Inclusion of pubertal stage did not modify findings and, given its correlation with age, it was not included in our final models.

### 1.5.5 Statistical Analysis

Statistical analyses were performed with SAS V.9.4. Mean differences in demographic variables between maltreatment and comparison groups were assessed via two-tailed t-tests for continuous variables and two-way Chi-Square tests for dichotomous variables. There were no statistically significant differences in demographic measures between the current total Time 1 cohort and the sub-sample included in the current analyses. Three individuals were missing covariate data on family income. No significant differences in demographic characteristics were found for these individuals, thus missing data were addressed simultaneously using multiple imputation and complete case analysis. We created 20 imputed datasets using PROC MI and combined imputed results with PROC MIANALYZE. Data were analyzed both with and without imputation. Imputed and complete-case datasets produced similar results with no changes in direction or size of effects, and we report results with the imputed data.

We assessed outcome variables for skewness via Box Cox analyses that indicated the marginal utility of a Y-1 transformation to the BMI and accelerated epigenetic aging variables. After running all models using both transformed and non-transformed outcome variables, we found no differences in the direction or significance of any findings and thus retained the non-transformed outcome models for ease of interpretability. All covariates survived assessment for multicollinearity via variance inflation factor analysis in PROC REG (OPTIONS = VIF and COLLINOINT) and were thus retained.

Though the majority of children included in our models were from unique families, siblings were included in the study and for our analytic sample we included four families with three siblings, 38 families with two siblings, and 185 families with individual children. To account for the partial-nesting of children within families (e.g., the violation of independence of child-level observations), all models were estimated with family-level cluster-robust standard errors in PROC GENMOD, with family ID as the repeated subject. Statistical significance was set at two-tailed *P*<.*05*. Unless specified, all data are presented as estimate (SE) with 95% confidence intervals. Where appropriate, models were corrected for multiple comparisons using Bonferroni adjusted p-values.

## Data Availability

The dataset analyzed during the current study is available through agreement with the Study investigators.

## Abbreviations

CM: Child Maltreatment
CWIS: Child Welfare Information System
CHS: Child Health Study
BMI: Body Mass Index
AA: Accelerated Aging
DunedinPoAm: Dunedin Pace of Aging methylation

## Acknowledgements

We thank the children and caregivers for their participation in the study, and Child Health Study staff for their dedication, hard work and insights. Research reported in this manuscript was supported by grants from the National Institutes of Health, National Institute of Child Health and Human Development P50HD089922 (J.G.N), R01HD072468 (J.G.N), the National Institutes of Aging through R01AG04879 (J.G.N). L.E. was supported by National Institute on Aging T32 AG049676 to The Pennsylvania State University. The content is solely the responsibility of the authors and does not necessarily represent the official views of the National Institutes of Health.

---

STROBE Statement—Checklist of items that should be included in reports of ***cohort studies***

|  | Item No | Recommendation | Page No |
| --- | --- | --- | --- |
| **Title and abstract** | 1 | (a) Indicate the study's design with a commonly used term in the title or the abstract<br>(b) Provide in the abstract an informative and balanced summary of what was done and what was found | 1 |
| **Introduction** |  |  |  |
| Background/rationale | 2 | Explain the scientific background and rationale for the investigation being reported | 3 |
| Objectives | 3 | State specific objectives, including any prespecified hypotheses | 5 |
| **Methods** |  |  |  |
| Study design | 4 | Present key elements of study design early in the paper | 5 |
| Setting | 5 | Describe the setting, locations, and relevant dates, including periods of recruitment, exposure, follow-up, and data collection | 11-12 |
| Participants | 6 | (a) Give the eligibility criteria, and the sources and methods of selection of participants. Describe methods of follow-up<br>(b) For matched studies, give matching criteria and number of exposed and unexposed | 12 |
| Variables | 7 | Clearly define all outcomes, exposures, predictors, potential confounders, and effect modifiers. Give diagnostic criteria, if applicable | 13-14 |
| Data sources/<br>measurement | 8* | For each variable of interest, give sources of data and details of methods of assessment (measurement). Describe comparability of assessment methods if there is more than one group | 13-14 |
| Bias | 9 | Describe any efforts to address potential sources of bias | 14-15 |
| Study size | 10 | Explain how the study size was arrived at | 11 |
| Quantitative variables | 11 | Explain how quantitative variables were handled in the analyses. If applicable, describe which groupings were chosen and why | 11-15 |
| Statistical methods | 12 | (a) Describe all statistical methods, including those used to control for confounding<br>(b) Describe any methods used to examine subgroups and interactions<br>(c) Explain how missing data were addressed<br>(d) If applicable, explain how loss to follow-up was addressed<br>(e) Describe any sensitivity analyses | 15-17 |
| **Results** |  |  |  |
| Participants | 13* | (a) Report numbers of individuals at each stage of study—eg numbers potentially eligible, examined for eligibility, confirmed eligible, included in the study, completing follow-up, and analysed (b) Give reasons for non-participation at each stage<br>(c) Consider use of a flow diagram | 11 |
| Descriptive data | 14* | (a) Give characteristics of study participants (eg demographic, clinical, social) and information on exposures and potential confounders<br>(b) Indicate number of participants with missing data for each variable of interest<br>(c) Summarise follow-up time (eg, average and total amount) | 5, T1, T S1 |
| Outcome data | 15* | Report numbers of outcome events or summary measures over time | 5, T1, T S1 |
| Main results | 16 | (a) Give unadjusted estimates and, if applicable, confounder-adjusted estimates and their precision (eg, 95% confidence interval). Make clear which confounders were adjusted for and why they were included<br>(b) Report category boundaries when continuous variables were categorized<br>(c) If relevant, consider translating estimates of relative risk into absolute risk for a meaningful time period | 5-7 |
| Other analyses | 17 | Report other analyses done—eg analyses of subgroups and interactions, and sensitivity analyses | Additional File |
| **Discussion** |  |  |  |
| Key results | 18 | Summarise key results with reference to study objectives | 5-6 |
| Limitations | 19 | Discuss limitations of the study, taking into account sources of potential bias or imprecision. Discuss both direction and magnitude of any potential bias | 6-7 |
| Interpretation | 20 | Give a cautious overall interpretation of results considering objectives, limitations, multiplicity of analyses, results from similar studies, and other relevant evidence | 5-7 |
| Generalisability | 21 | Discuss the generalisability (external validity) of the study results | 6-8 |
| **Other information** |  |  |  |
| Funding | 22 | Give the source of funding and the role of the funders for the present study and, if applicable, for the original study on which the present article is based | 18 |

\*Give information separately for exposed and unexposed groups.
**Note:** An Explanation and Elaboration article discusses each checklist item and gives methodological background and published examples of transparent reporting. The STROBE checklist is best used in conjunction with this article (freely available on the Web sites of PLoS Medicine at http://www.plosmedicine.org/, Annals of Internal Medicine at http://www.annals.org/, and Epidemiology at http://www.epidem.com/). Information on the STROBE Initiative is available at http://www.strobe-statement.org.

