## Supplemental Tables for "Obesity and accelerated epigenetic aging in a high-risk cohort of children"

**Additional File**

**Tables.**

Supplementary Table 1. Summary Statistics for Currently Assembled Cohort

| **Variable, mean (SD)** | **Total (N=439)** | **Maltreatment (N=373)** | **Comparison (N=66)** | **P-value** |
|---|---|---|---|---|
| Body Mass Index (BMI) | 21.6(5.8) | 22.0(5.9) | 19.9(4.8) | .0009** |
| Age (at collection) | 11.4(1.4) | 11.5(1.4) | 11.1(1.4) | .08 |
| Biological Sex M(F) | 51.5%(48.5) | 51.5%(48.5) | 51.5%(48.5) | .99 |
| Income $10,000/year | 3.7(3.3) | 3.3(3.1) | 5.8(3.8) | <.0001*** |
| Race | | | | |
| Black | 15.0% | 15.6% | 12.1% | .47 |
| White | 71.3% | 70.2% | 77.3% | .24 |
| Other | 13.7% | 14.2% | 11.7% | .43 |
| Ethnicity | | | | |
| Hispanic | 13.2% | 14.8% | 4.6% | .02* |
| *†P<.10, *P<.05, **P<.01, ***P<.0001* | | | | |

Supplementary Table 2. HorvathAA predicted by BMI

| Outcome: HorvathAA | Hypothesis 1a | | | | | Hypothesis 1b | |
|---|---|---|---|---|---|---|---|
| | Model 1 | Model 2 | Model 3 | Model 4 | Model 5 | Model 6a | Model 6b |
| | Est.±SE | Est.±SE | Est.±SE | Est.±SE | Est.±SE | Est.±SE | Est.±SE |
| Parameter | | | | | | | |
| BMI | 0.02(.02) | 0.02(.02) | 0.02(.02) | 0.02(.02) | 0.02(.02) | 0.02(.02) | 0.06(.06) |
| Sex (Male) | -- | 0.26(.24) | 0.26(.25) | 0.26(.25) | 0.21(.25) | 0.21(.25) | 0.20(.25) |
| Race/Ethnicity (Ref=white/non-Hispanic) | | | | | | | |
| Black | -- | -- | -0.02(.35) | -0.10(.37) | 0.26(.37) | 0.26(.36) | 0.29(.37) |
| Other | -- | -- | 0.26(.37) | 0.22(.38) | 0.35(.35) | 0.34(.35) | 0.35(.35) |
| Hispanic | -- | -- | -0.36(.43) | -0.41(.44) | -0.26(.39) | -0.26(.40) | -0.24(.40) |
| Household Income | -- | -- | -- | -0.03(.04) | -0.02(.04) | -0.02(.04) | -0.02(.04) |
| Proportion Lymphocytes | -- | -- | -- | -- | -0.67(.17)** | -0.67(.17)** | -0.69(.17)*** |
| Proportion Monocytes | -- | -- | -- | -- | -0.76(.19)*** | -0.76(.19)*** | -0.78(.19)*** |
| Proportion Granulocytes | -- | -- | -- | -- | -0.69(.18)*** | -0.68(.18)** | -0.70(.18)** |
| Maltreatment | -- | -- | -- | -- | -- | 0.06(.33) | 0.94(1.26) |
| BMI X Maltreatment | -- | -- | -- | -- | -- | -- | -0.04(.06) |

*† P<.10, *P<.05, **P<.01, ***P<.0001*

Supplementary Table 3. HannumAA predicted by BMI

| Outcome: HannumAA | Hypothesis 1a | | | | | Hypothesis 1b | |
|---|---|---|---|---|---|---|---|
| | Model 1 | Model 2 | Model 3 | Model 4 | Model 5 | Model 6a | Model 6b |
| | Est.±SE | Est.±SE | Est.±SE | Est.±SE | Est.±SE | Est.±SE | Est.±SE |
| Parameter | | | | | | | |
| BMI | 0.06(.03)* | 0.06(.03)† | 0.05(.03)† | 0.05(.03)† | -0.03(.03) | -0.03(.03) | 0.04(.07) |
| Sex (Male) | -- | -0.44(.38) | -0.34(.38) | -0.34(.38) | -0.13(.31) | -0.12(.31) | -0.13(.31) |
| Race/Ethnicity (Ref=white/non-Hispanic) | | | | | | | |
| Black | -- | -- | -0.62.57) | -0.70(.60) | 0.37(.45) | 0.36(.45) | 0.41(.46) |
| Other | -- | -- | -0.02(.58) | -0.07(.59) | 0.39(.43) | 0.42(.44) | 0.43(.44) |
| Hispanic | -- | -- | 1.09(.48)* | 1.04(.50)* | 1.33(.49)** | 1.34(.49)** | 1.38(.49)** |
| Household Income | -- | -- | -- | -0.02(.06) | 0.03(.05) | 0.02(.05) | 0.02(.05) |
| Proportion Lymphocytes | -- | -- | -- | -- | -0.98(.22)*** | -0.97(.22)*** | -1.01(.22)*** |
| Proportion Monocytes | -- | -- | -- | -- | -0.73(.23)** | -0.73(.23)** | -0.76(.23)** |
| Proportion Granulocytes | -- | -- | -- | -- | -0.86(.23)** | -0.86(.23)** | -0.89(.23)*** |
| Maltreatment | -- | -- | -- | -- | -- | -0.27(.41) | 1.37(1.57) |
| BMI X Maltreatment | -- | -- | -- | -- | -- | -- | -0.08(.08) |

*† P<.10, *P<.05, **P<.01, ***P<.0001*

Supplementary Table 4. GrimAgeAA predicted by BMI

| Outcome: GrimAgeAA | Hypothesis 1a | | | | | Hypothesis 1b | |
|---|---|---|---|---|---|---|---|
| | Model 1 | Model 2 | Model 3 | Model 4 | Model 5 | Model 6a | Model 6b |
| | Est.±SE | Est.±SE | Est.±SE | Est.±SE | Est.±SE | Est.±SE | Est.±SE |
| Parameter | | | | | | | |
| BMI | 0.12(.02)*** | 0.13(.02)*** | 0.12(.02)*** | 0.11(.02)*** | 0.06(.02)** | 0.05(.02)** | 0.15(.05)** |
| Sex (Male) | -- | 0.76(.27)** | 0.74(.28)** | 0.74(.26)** | 0.92(.25)** | 0.91(.25)** | 0.89(.25)** |
| Race/Ethnicity (Ref=white/non-Hispanic) | | | | | | | |
| Black | -- | -- | 1.04(.57)† | 0.50(.58) | 1.04(.36) | 1.05(.36)** | 1.12(.36)** |
| Other | -- | -- | 0.05(.41) | -0.25(.38) | -0.05(.35) | -0.09(.35) | -0.07(.35) |
| Hispanic | -- | -- | 0.34(.40) | -0.004(.37) | 0.14(.39) | 0.12(.39) | 0.17(.39) |
| Household Income | -- | -- | -- | -0.19(.05)** | -0.16(.04)*** | -0.15(.04)** | -0.14(.04)** |
| Proportion Lymphocytes | -- | -- | -- | -- | -0.37(.17)* | -0.38(.17)* | -0.43(.17)* |
| Proportion Monocytes | -- | -- | -- | -- | -0.25(.19) | -0.25(.19) | -0.30(.19) |
| Proportion Granulocytes | -- | -- | -- | -- | -0.29(.18) | -0.30(.18)† | -0.34(.18)† |
| Maltreatment | -- | -- | -- | -- | -- | 0.44(.32) | 2.68(1.25)* |
| BMI X Maltreatment | -- | -- | -- | -- | -- | -- | -0.11(.06)† |

*† P<.10, *P<.05, **P<.01, ***P<.0001*

Supplementary Table 5. PhenoAgeAA predicted by BMI

| Outcome: PhenoAgeAA | Hypothesis 1a | | | | | Hypothesis 1b | |
|---|---|---|---|---|---|---|---|
| | Model 1 | Model 2 | Model 3 | Model 4 | Model 5 | Model 6a | Model 6b |
| | Est.±SE | Est.±SE | Est.±SE | Est.±SE | Est.±SE | Est.±SE | Est.±SE |
| Parameter | | | | | | | |
| BMI | 0.25(.06)*** | 0.23(.06)** | 0.23(.06)** | 0.22(.06)** | 0.05(.04) | 0.05(.04) | 0.07(.11) |
| Sex (Male) | -- | -2.30(.69)** | -2.28(.69)** | -2.28(.69)** | -1.56(.51)** | -1.56(.51)** | -1.57(.52)** |
| Race/Ethnicity (Ref=white/non-Hispanic) | | | | | | | |
| Black | -- | -- | -0.82(.94) | -1.14(.96) | 0.71(.75) | 0.72(.75) | 0.74(.75) |
| Other | -- | -- | 0.12(1.11) | -0.06(1.10) | 0.54(.72) | 0.53(.72) | 0.53(.72) |
| Hispanic | -- | -- | -0.38(1.06) | -0.58(1.08) | -0.03(.81) | -0.05(.81) | -0.03(.82) |
| Household Income | -- | -- | -- | -0.11(.11) | 0.003(.08) | 0.01(.08) | 0.01(.08) |
| Proportion Lymphocytes | -- | -- | -- | -- | -1.26(.36)** | -1.26(.36)** | -1.27(.36)** |
| Proportion Monocytes | -- | -- | -- | -- | -0.93(.38)* | -0.93(.38)* | -0.94(.39)* |
| Proportion Granulocytes | -- | -- | -- | -- | -0.94(.37)* | -0.94(.37)* | -0.96(.37)* |
| Maltreatment | -- | -- | -- | -- | -- | 0.21(.67) | 0.77(2.59) |
| BMI X Maltreatment | -- | -- | -- | -- | -- | -- | -0.03(.12) |

*† P<.10, *P<.05, **P<.01, ***P<.0001*

Supplementary Table 6. DunedinPoAm predicted by BMI

| Outcome: DunedinPoAm | Hypothesis 1a | | | | | | Hypothesis 1b | |
|---|---|---|---|---|---|---|---|---|
| | Model 1 | Model 2 | Model 3 | Model 4 | Model 5 | Model 6 | Model 7a | Model 7b |
| | Est.±SE | Est.±SE | Est.±SE | Est.±SE | Est.±SE | Est.±SE | Est.±SE | Est.±SE |
| Parameter | | | | | | | | |
| BMI | 0.003(.0004)*** | 0.003(.0004)*** | 0.003(.0004)*** | 0.003(.0004)*** | 0.003(.0004)*** | 0.002(.0004)*** | 0.002(.0004)*** | 0.002(.0009)* |
| Sex (Male) | -- | -0.01(.005)† | -0.01(.005)† | -0.01(.005)† | -0.01(.005)† | -0.004(.004) | -0.004(.004) | -0.004(.004) |
| Age | -- | -- | -0.004(.002)* | -0.004(.002)* | -0.004(.002)* | -0.004(.001)** | -0.004(.002)** | -0.004(.001)** |
| Race/Ethnicity (Ref=white/non-Hispanic) | | | | | | | | |
| Black | -- | -- | -- | 0.006(.006) | 0.004(.007) | 0.02(.006)* | 0.02(.006)* | 0.02(.006)* |
| Other | -- | -- | -- | 0.006(.008) | 0.005(.008) | 0.008(.006) | 0.008(.006) | 0.008(.006) |
| Hispanic | -- | -- | -- | -0.0001(.008) | -0.001(.008) | 0.002(.007) | 0.002(.007) | 0.002(.007) |
| Household Income | -- | -- | -- | -- | -0.001(.001) | 0.0002(.0006) | 0.0004(.0006) | 0.0004(.0006) |
| Proportion Lymphocytes | -- | -- | -- | -- | -- | -0.005(.002)† | -0.005(.003)† | -0.005(.003)† |
| Proportion Monocytes | -- | -- | -- | -- | -- | -0.003(.003) | -0.003(.003) | -0.003(.003) |
| Proportion Granulocytes | -- | -- | -- | -- | -- | -0.003(.003) | -0.003(.003) | -0.003(.003) |
| Maltreatment | -- | -- | -- | -- | -- | -- | 0.007(.005) | 0.01(.02) |
| BMI X Maltreatment | -- | -- | -- | -- | -- | -- | -- | -0.0002(.001) |

† *P<.10, *P<.05, **P<.01, ***P<.0001*

Supplementary Table 7. BMI predicted by maltreatment-status

| Outcome: BMI | Model 1 | Model 2 | Model 3 | Model 4 | Model 5 |
|---|---|---|---|---|---|
| | Est.±SE | Est.±SE | Est.±SE | Est.±SE | Est.±SE |
| Parameter | | | | | |
| Maltreatment | 1.78(.96) | 1.41(.94) | 1.41(.93) | 1.30(.94) | 0.70(.97) |
| Age | -- | 1.13(.25)*** | 1.11(.25)*** | 1.07(.24)*** | 1.10(.23)*** |
| Sex (Male) | -- | -- | -1.21(.69)† | -1.12(.70) | -1.08(.69) |
| Race/Ethnicity (Ref=white/non-Hispanic) | | | | | |
| Black | -- | -- | -- | 0.24(.88) | -0.59(.95) |
| Other | -- | -- | -- | 0.001(1.08) | -0.37(1.09) |
| Hispanic | -- | -- | -- | 1.35(1.39) | 0.82(1.45) |
| Household Income | -- | -- | -- | -- | -0.30(.11)** |

† *P<.10, *P<.05, **P<.01, ***P<.0001*
